# Immune response to SARS-CoV-2 mRNA vaccination in multiple sclerosis patients after rituximab treatment interruption

**DOI:** 10.1101/2023.02.21.23286229

**Authors:** Remigius Gröning, Andy Dernstedt, Clas Ahlm, Johan Normark, Peter Sundström, Mattias NE Forsell

## Abstract

Peripheral B cell depletion via anti-CD20 treatment is a highly effective disease-modifying treatment in multiple sclerosis (MS) patients. A drawback of anti-CD20 treatment is poor immune responses to vaccination. While this can be mitigated by treatment interruption of at least six months prior to vaccination, the timing to resume treatment while maintaining subsequent vaccine responses remains undetermined. We characterized SARS-CoV-2 S-directed antibody and B cell responses throughout three BNT162b2 doses in MS patients, where the first two doses were given during treatment interruption. The last anti-CD20 rituximab infusion was given 1.3 years (median) prior to the first vaccine dose and re-administered four weeks after the second vaccine dose. After two vaccine doses, antibody-mediated responses in SARS-CoV-2-naïve MS patients were comparable to vaccinated healthy controls, albeit with greater variation. We could demonstrate that the response to the second dose of vaccination was predictive of a boost effect after a third dose, even after re-initiation of rituximab. MS patients also exhibited lower frequencies of Decay Accelerating Factor-negative memory B cells, a suggested proxy for germinal centre activity, than healthy individuals. Our findings also offer a first indication on the potential importance of antigenic stimulation of CD27^-^IgD^-^ double negative B cells and the possible long-term impairment of germinal centre activity in rituximab-treated MS patients.

## Introduction

Multiple sclerosis (MS) is an autoimmune disease that causes inflammation of the central nervous system^1^. Although the underlying cause remains unknown, disease susceptibility is increased through a combination of heritable and environmental factors^2^. Traditional disease modifying treatments (DMT), such as broad immunosuppressive and –modulatory drugs have a positive impact on the relapse rate of MS. However, their effect on disability advancement is limited during progressive MS^3^. The suspected involvement of B cells in MS pathophysiology was proven in experimental trials using treatments specifically targeting B cells^1,4^. Rituximab (RTX) is a genetically engineered chimeric monoclonal antibody that depletes CD20^+^ cells, including most B cell subsets except CD20^-^ plasma cells. Accordingly, RTX has been shown to be highly effective to reduce inflammatory brain lesions and clinical relapses for a prolonged period^4,5^. Therefore, the treatment has been introduced as standard-of-care for MS patients at many hospitals in Sweden and elsewhere^6^. The inhibition of the B cell-dependent adaptive immune system by RTX and other DMTs has also been shown to lead to a higher rate of hospitalization due to COVID-19^7-10^.

Due to its B cell-depleting effect, RTX treatment is detrimental for vaccine-induced responses. Treatment interruption is therefore required to mount efficient vaccine-induced responses, as has been shown^11,12^. In the case of MS patients, non-live COVID-19 vaccines, like the mRNA vaccine BNT162b2, were demonstrated safe and effective^13^. At Umeå University Hospital, MS patients were under RTX treatment interruption prior to the first dose of BNT162b2, then resumed RTX approximately four weeks after the second vaccine dose. Here we studied B cell responses to repeated vaccination with BNT162b2 during RTX treatment interruption, with the goal to identify factors that could predict the efficacy of booster vaccinations after RTX treatment re-initiation.

## Patients and methods

### Patient cohort and material

The study was approved by the Swedish Ethics Review Authority (Dnr 2021-00055, including approved amendments) and the Medical Products Agency Sweden. The study was registered at European Clinical Trials Database (EUDRACT Number 2021-000683-30) before the first patient was enrolled in the study. Umeå University, Sweden served as trial sponsor and the Clinical Research Center, University Hospital of Northern Sweden was monitoring the study for regulatory compliance. All individuals were included after informed consent and data were stored in accordance with the EU General Data Protection Regulation. We enrolled 43 MS patients via the Neurological Department at Umeå University Hospital after informed consent, of which 42 had sufficient samples drawn for a longitudinal study (Table 1, Figure 1). All had relapsing-remitting disease and were at present exclusively treated with RTX. Eight out of 43 individuals (19 %) contracted COVID-19 before vaccination. All were vaccinated with Pfizer-BioNTech’s BNT162b2 mRNA vaccine half to two-and-a-half years after their last RTX infusion. The control cohort comprised 20 age- and sex-matched healthy controls, that were vaccinated with BNT162b2 (Table 1). For the antibody avidity assay, we studied a different control cohort consisting of 10 individuals, which received three doses of BNT162b2.

**Table 1.** Clinical characteristics of 43 relapsing-remitting multiple sclerosis cases treated with RTX and 20 non-MS control individuals.

| Characteristics | All patients (N=43) | Non-MS control group (N=20) |
| --- | --- | --- |
| Female n (%) | 26 (60) | 12 (60) |
| Age at disease onset | 26 (15-47) |  |
| Years of disease duration | 9 (1-25) |  |
| Age at enrollment | 38 (25–64) | 41.5 (20-59) |
| BMI | 25 (19-43) | 23.6 (18.7-41.6) |
| EDSS | 1.0 (0-4.0) |  |
| Covid 19 before vaccination n | 8 | 0 |
| **Clinical parameters\*** |  |  |
| B cell count (cells/μl) | 123 (1-817) | |
| Vitamin D level (nmol/l) | 85 (54-149) |  |
| Lymphocyte count ($10^9$/l) | 1.9 (1-3.4) | |
| CRP (mg/ml) | 1.3 (0.6-20) |  |
| Comorbidities | None (n=29), asthma (n=7), hypertension (n=2), hypothyroidism (n=2), diabetes type II (n=1), obesity (n=1), hyperlipidemia (n=1), | None (n=5), allergy (n=1), asthma (n=5), vaso-occlusive crisis (n=2), diabetes type I (n=1), gastritis (n=1), IgA nephritis (n=1), migraine (n=2) |
|  | prolactinoma (n=1), activated<br>protein C resistance (n=1) |  |
| **Rituximab treatment** |  |  |
| Number of rituximab infusions<br>before vaccine dose 1 | 8 (1-11) |  |
| Rituximab total dosage (mg)<br>before vaccine dose 1 | 5500 (1000-10500) |  |
| Rituximab last infusion dose (mg) | 500 (300-1000) |  |
| Years since last rituximab infusion<br>at vaccine dose 1 | 1.3 (0.52-2.6) |  |
| **DMD before rituximab n (%)** |  |  |
| None | 19 (44) |  |
| Interferon beta | 19 (44) |  |
| Natalizumab | 14 (33) |  |
| Dimethyl fumarate | 1 (2.3) |  |
| Glatiramer acetate | 2 (4.7) |  |
| Fingolimod | 3 (7.0) |  |

All figures are median (range) unless stated otherwise. BMI, body mass index; CRP, c-reactive protein; DMD, disease modifying drug; EDSS, Extended Disability Status Scale.
\*B cell (CD19+) count, vitamin D levels (25[OH]D), lymphocyte count, and CRP from the latest sample preceding rituximab treatment re-initiation.

**Figure 1:**
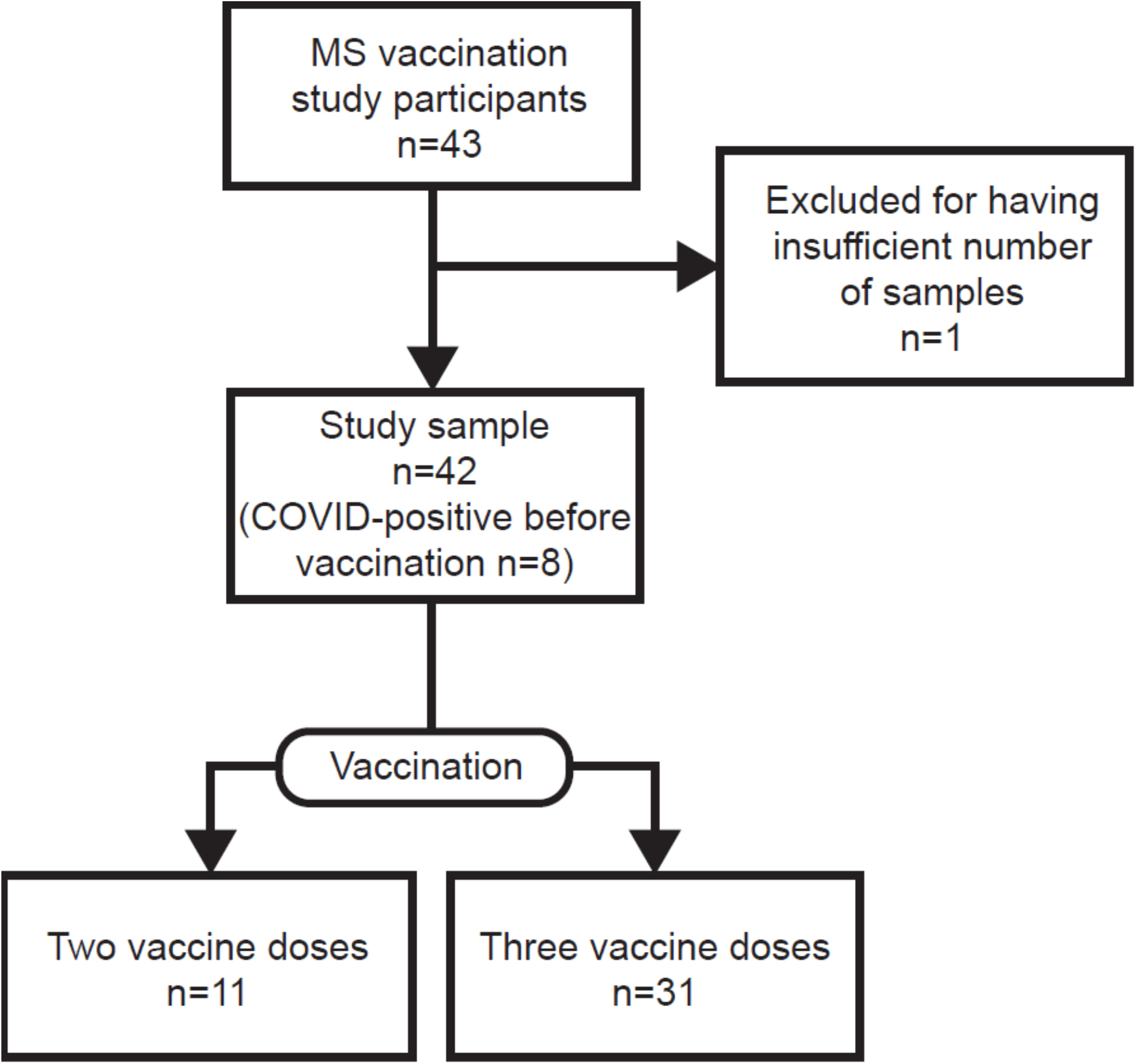
Inclusion flowchart of study cohort. Depicted are the number of vaccinated rituximab-treated (RTX) multiple sclerosis (MS) patients participating in the study and the final study sample after exclusion. The study sample is characterized further by the number of vaccine doses and COVID-experienced participants.

According to a consensus document by the Swedish MS-association regarding general vaccinations for MS patients on DMTs^14^, RTX retreatment was postponed until one month after two BNT162b2 doses, with the first dose given when the B cell count exceeded 20 cells/µl and the second subsequently one month after. Thus, RTX treatment resumed before the third vaccine dose.

Specific blood sampling numbers and intervals are described in supplemental Method 1 and supplemental Figure 1. Blood was collected in sodium-heparin CPT tubes (BD, 362780) and serum gel tubes (BD, 366566) for the isolation of peripheral blood mononuclear cells (PBMCs) and serum, respectively, according to manufacturer’s recommendations. PBMCs were cryopreserved in FBS supplemented with 10% DMSO and stored in liquid nitrogen. Lymphocyte count, flow cytometry B-cell count (CD19^+^ cells), 25(OH)D, and c-reactive protein (CRP) routinely analyzed at the hospital laboratory were retrieved from the last blood sample before the first vaccine dose. The last estimate of MS disability was assessed using Extended Disability Status Scale (EDSS)^15^.

The study was part of a clinical trial to study immunogenicity of COVID-19 vaccination (EudraCT 2021-000683-30, ClinicalTrials.gov ID NCT04920357). Ethical approvement was obtained by the Swedish Ethical Review Authority (Dnr 2021-00055 and subsequent approved amendments).

### Production of SARS-CoV-2-S protein and -pseudotyped lentivirus

The 2019-nCoV S protein was produced using the Gibco ExpiCHO Expression System (Thermo Fisher Scientific) and purified as previously described in Byström *et al*.^16^.

A lentivirus containing the luciferase reporter gene and 2019-nCoV-D614G-S protein was produced with the FuGENE 6 transfection reagent (Promega) according to the manufacturer’s protocol. See supplemental Method 2 for a detailed description.

### Titration of pseudotyped lentivirus

293T-hACE2.MF cells (provided by Nicole Doria-Rose) were seeded at a density of 5000 cells per well in a 96-well black cell culture plate (Greiner) in Dulbecco’s Modified Eagle Medium (DMEM) (Thermo Scientific) + 10 % fetal bovine serum (FBS) + 1000 U/L Penicillin+Streptomycin (D10 medium) and incubated at 37 °C, 5% CO2. On the next day, the pseudotyped lentivirus was thawed and diluted in a 96-well deep plate (Corning) in the following manner. 1:2 dilutions of pseudovirus with Minimum Essential Medium (MEM, Thermo Scientific) + 5 % FBS (MEM5 medium) were prepared in the 11 wells of the first row. The last well contained only MEM5 medium. The dilution plate was incubated for 45 min at 37 °C. The medium from the cells prepared on the previous day was removed and 50 µL of pseudovirus dilution was added to the cells in 8 replicates and incubated for 2 h at 37 °C, 5% CO2. Afterwards, 150 µL of MEM5 medium was added to each well and incubated for 3 days at 37 °C, 5% CO2.

After 3 days, cells were lysed, and luciferase reagent added according to the manufacturer’s instructions (Luciferase Assay System, Promega). The luminescence was measured on a Tecan Sunrise microplate reader within 2 minutes of adding the reagent with 1 second integration time. A pseudovirus dilution step for the neutralization assay was selected that yielded 1000-times the luminescence value of the medium-only wells.

### Neutralization assay

Neutralization of the authentic SARS-CoV-2 virus was conducted as previously described in Byström *et al*.^16^. For the neutralization of the pseudotyped lentivirus, serum samples were inactivated for 20 min at 56 °C. 5000 cells of 293T-hACE2.MF per well were seeded in a 96-well black cell culture plate (Greiner) in D10 medium and incubated at 37 °C, 5% CO2. The next day, for each serum sample an 8-point 3-fold dilution series of the sample was prepared in a V-bottom 96-well plate in MEM5 medium. Then, pseudovirus diluted at half the chosen dilution was added to each serum dilution point at 1:1 ratio. Four wells per cell plate contained MEM5 medium, as well as four wells with the chosen pseudovirus dilution. The dilution plate was incubated for 45 min at 37 °C. Afterwards, the medium was removed from the cells and 50 µL of the serum sample-pseudovirus mix was added in duplicates and incubated for 2 h at 37 °C, 5% CO2. Afterwards, 150 µL of M5 medium was added to each well and incubated for 3 days at 37 °C, 5% CO2. After 3 days the plates were prepared and measured as described in the previous paragraph.

### SARS-CoV-2 spike-specific enzyme-linked immunosorbent assay (ELISA) and avidity assay

The ELISA was conducted as previously described in Byström *et al*.^16^, with patient serum samples diluted 1/50 in blocking buffer, and seven additional 5-fold serial dilutions.

For the avidity assay, the dilution series was prepared in duplicates. Before the addition of the conjugate antibody, the second replicate of the dilution series was incubated with 1.5 M NaSCN in PBS for 10 min at RT and the plates subsequently washed. The avidity index was determined by the AUC of the NaSCN-treated replicate divided by the AUC of the untreated replicate.

### Flow cytometric characterisation of B cells

Isolated Purified SARS-CoV-2 spike protein was tetramerised with Strep-Tactin conjugated to APC and PE (IBA), respectively, for a minimum of 20 minutes at 4°C protected from light. Cryopreserved PBMCs were thawed in a 37°C water bath, washed in sterile PBS supplemented with 2% FBS (PBS+FBS), and transferred to a V-bottom 96-well plate for staining. Unless otherwise specified, all staining steps of the cells were performed at 4°C protected from light. 25nM each of APC and PE conjugated protein were incubated on the PBMCs for 60 mins. Further, cells were stained with Zombie NIR (Biolegend, 1:1000) according to manufacturer’s instructions. Antibodies targeted towards surface antigens (supplemental Table 1) were diluted in PBS+FBS and incubated for 25 min. Cells were fixed with eBioscience FoxP3/Transcription Factor Staining Buffer Set (Thermo Scientific), according to manufacturer’s instructions. Finally, cells were resuspended in sterile PBS+FBS and strained through 70µm pre-separation filters (Miltenyi) before acquisition on a BioRad ZE5 flow cytometer. The acquisition speed was 1 µl/second and sample temperature 4°C during acquisition. FCS files were analysed using FlowJo v10. Total B cells were defined as viable CD19^+^CD20^+^, and memory populations were distinguished by their expression of CD27 and IgD.^17,18^

### Statistical analysis

For the neutralization assay, all values were subtracted by the average value of the medium-only wells and divided by the average of pseudovirus-only wells. The serum dilution required to reduce the pseudovirus infection by 50% (ID50) was determined with GraphPad Prism 9 (GraphPad Software). For the ELISA, all values were subtracted by the average value of the blocking buffer-only wells. Area under the curve (AUC) of the dilution curve was determined with GraphPad Prism 9 (GraphPad Software), with a baseline level of 0.2.

A mixed model with Geisser-Greenhouse correction and matched timepoints per individual was used together with Tukey’s multiple comparisons test for comparing S-binding IgG and neutralizing antibody levels between separate groups and time points. A Spearman correlation test was used for correlation analysis of multiple serology, clinical, and demographic data. All analysis was done in Graphpad Prism 9 (GraphPad Software). Chosen parameters for Spearman correlation are listed in supplemental Method 3. For the flow cytometry results, comparisons within groups were done by Wilcoxon matched-pairs signed rank test, and comparisons between groups by Mann-Whitney’s test, using Graphpad Prism 9 (GraphPad Software).

## Results

### Higher titers of neutralizing antibodies in COVID-experienced than COVID-naïve MS patients after vaccination

Of the 43 study participants in the MS group, we were able to acquire longitudinal samples from 42 individuals (Figure 1). Of these, all had been diagnosed with relapsing-remitting MS and been on RTX treatment interruption for a median of 1.3 years prior to BNT162b2 vaccination (Table 1). This allowed for reconstitution of peripheral B cells above 20 cells/µL prior to receiving the 1^st^ vaccine dose. Additional demographic and clinical data for the MS patients included in this study are shown in Table 1. RTX treatment was resumed for all patients at four weeks after the second dose of BNT162b2. Eight out of 43 individuals (19%) contracted COVID-19 before vaccination but after treatment interruption. Blood sampling of patients in the context to the three vaccine doses is described in detail in supplemental Figure 1.

We assessed circulating SARS-CoV-2 binding (Figure 2A,B) and neutralizing (Figure 2C,D) antibody levels to evaluate the response to the BNT162b2 vaccine. We could demonstrate that two doses of BNT162b2 induced similar levels of S-binding antibodies in SARS-CoV-2 naïve patients as in healthy individuals, and that this similarity remained up until eight months after the second dose (Figure 2A). Consistently, two doses of BNT162b2 during treatment interruption induced similar levels of neutralizing antibodies against the SARS-CoV-2 S-pseudotyped or authentic virus in patients and in matched control subjects (Figure 2A, supplemental Figure 2). These data demonstrated that the time of treatment interruption had been sufficient to allow for efficient development of S-binding antibodies after a two-dose regimen of BNT162b2. In contrast, we found that the avidity of elicited S-binding IgG was lower in COVID-naïve patients than in healthy controls at one week (1.9-fold, p = 0.003), three months (1.6-fold), and eight months (1.8-fold) after two vaccine doses (Figure 2C).

**Figure 2:**
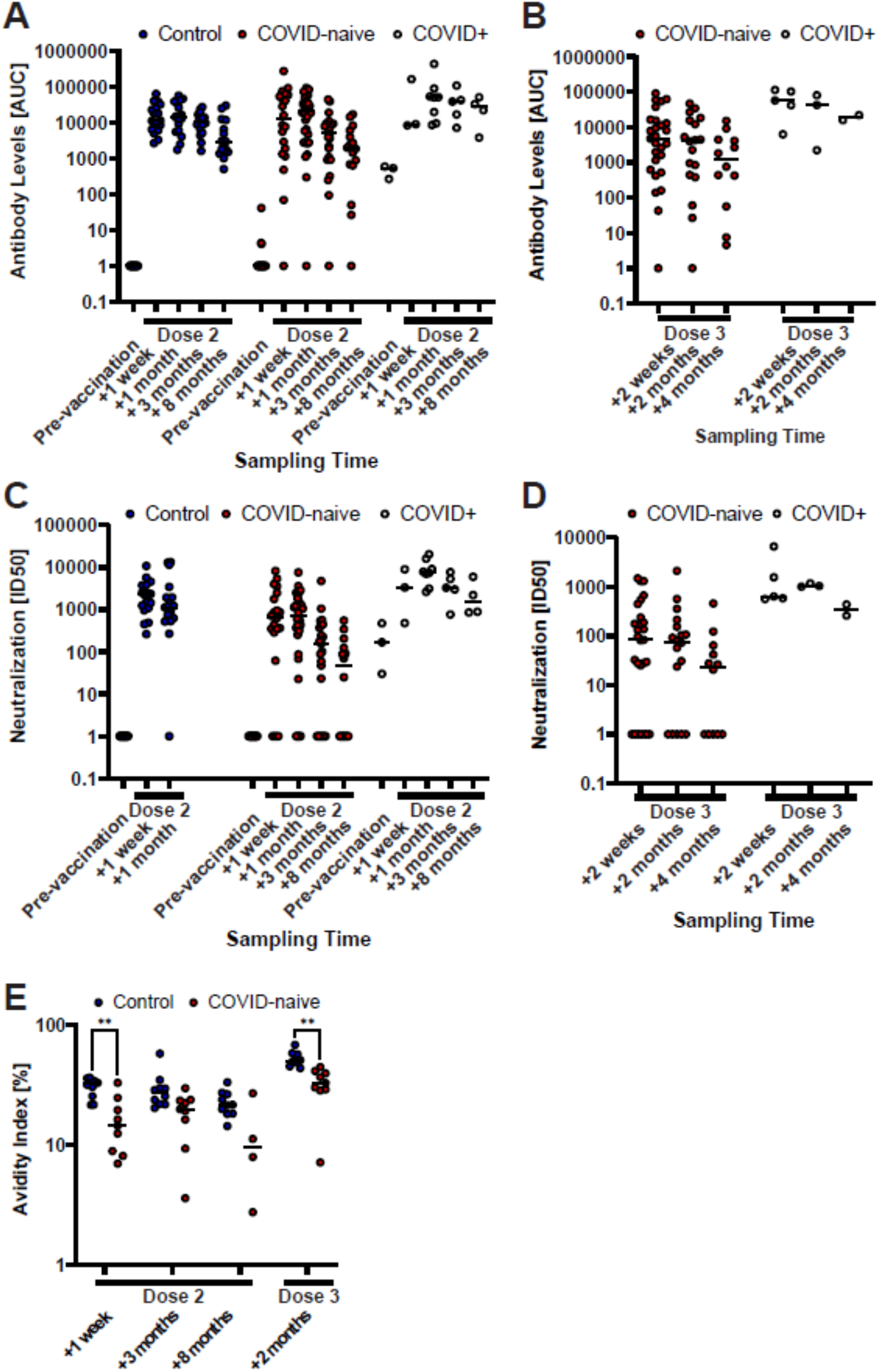
Generally impaired humoral immune response in COVID-naïve multiple sclerosis (MS) patients after vaccination. Longitudinal SARS-CoV-2 spike-specific antibody levels after two (A) and three (B) vaccine doses, and SARS-CoV-2-pseudotyped lentivirus neutralization capability after two (C) and three (D) vaccine doses of the non-MS control group and patient groups, the latter differentiated between patients that were infected with COVID-19 prior to vaccination (COVID+) and COVID-naïve patients. Antibody levels are described as the area under the curve (AUC) of a patient serum dilution series. Neutralization capability is described as the serum dilution required to reduce the viral infection by 50% (ID50). The horizontal, dashed line displays the reference value of a low-neutralizing control serum sample. Sample points, whose neutralizing capability or antibody levels were below detection have been set to 1 for illustrative purposes. (E) Longitudinal antibody avidity of a subset (n = 10) of the non-MS control and COVID-naïve patient groups. * p<0.05; ** p<0.01; *** p<0.001; **** p<0.0001.

Treatment with RTX was resumed for all patients four weeks after the second dose of BNT162b2. A third vaccine dose was given after an additional seven months. In contrast to the strong response in healthy individuals^17,18^, we found that a third dose in patients was inefficient to significantly boost antibody-mediated responses on a group level (Figure 2B,D). A third dose led to an increase in avidity of circulating S-specific IgG in both control and patient groups, but the patient’s IgG were of lower (1.6-fold, p = 0.001) avidity than those of the healthy controls (Figure 2E). Finally, patients who had been infected with SARS-CoV2 during RTX treatment interruption, but before vaccination, responded strongly to two doses of BNT162b2 (Figure 2A,C). Since we wanted to study the de-facto vaccine effect, COVID-experienced patients were excluded from further analysis.

### A low response to primary vaccination pre-disposes MS patients to low or absent response to a consecutive booster dose after continuation of RTX treatment

We noted that the variance of S-binding IgG was greater among patients compared to healthy controls. We therefore divided patients into two groups, low and high responders, based on the levels of circulating anti-S IgG after the third dose of BNT162b2. The median difference in levels of circulating anti-S IgG between the two subgroups was 36-fold (p = 0.007) at one week after the third dose. When analysing these individuals retrospectively, we found that the low responder group had consistently lower levels of anti-S IgG in circulation after the second dose of BNT162b2 (p = 0.04 for one week after second dose, p = 0.03 for three months after second dose, Figure 3A). This was also reflected in lower levels of circulating SARS-CoV-2 neutralizing antibodies throughout all time points, albeit not significantly (Figure 3B). Importantly, we found that the boost effect of the third dose (fold-change) was significantly lower in the low responder group (p = 0.006, Figure 3C).

**Figure 3:**
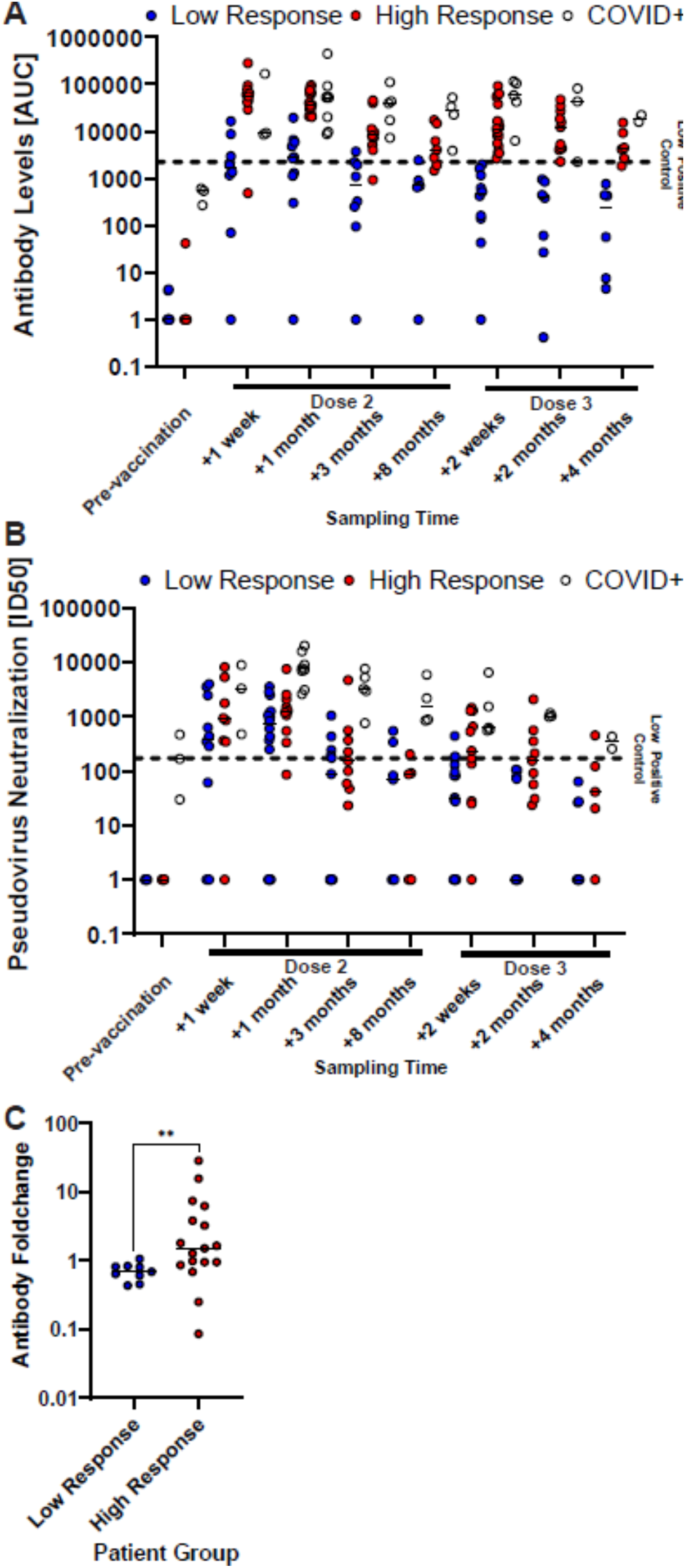
COVID-naïve multiple sclerosis (MS) patients with high SARS-CoV-2 antibody levels after the second vaccine dose showed a consistently high humoral immune response. SARS-CoV-2 spike-specific antibody levels (A) and SARS-CoV-2-pseudotyped lentivirus neutralization capability (B) of the patient groups, differentiating between patients that were infected with COVID-19 prior to vaccination (COVID+) and COVID naïve patients. COVID naïve patients were further grouped according to if their antibody levels were higher (High Response) or lower (Low Response) than the low-neutralizing control serum sample one week after the third vaccine dose. Antibody levels are described as the area under the curve (AUC) of a patient serum dilution series. Neutralization capability is described as the serum dilution required to reduce the viral infection by 50% (ID50). The horizontal, dashed line displays the reference value of a low-neutralizing control serum sample. Samples points, whose neutralizing capability or antibody levels were below detection have been set to 1 for illustrative purposes. (C) Anti-S IgG level foldchange between two weeks after and the last available time point (four to eight months) before the third vaccine dose of COVID-naïve MS patients.

### Characterization of vaccine-induced B cells in MS patients after RTX treatment interruption

We performed flow cytometric analysis of the circulating B cell compartment of patients and healthy control subjects to investigate if the low response in certain individuals could be due to differential B cell activation after treatment interruption and two BNT162b2 doses. Consistent with the long interval of RTX treatment interruption, we found a similar frequency of circulating total B cells (CD19^+^) in patients as in healthy controls up until four weeks post two BNT162b2 doses (Figure 4A,B). Accordingly, RTX treatment re-initiation four weeks after the second BNT162b2 dose led to efficient depletion of the peripheral B cell compartment in patients (Figure 4B). We then assessed the distribution of switched memory B cells (CD19^+^CD20^+^CD27^+^IgD^-^), unswitched memory B cells (CD19^+^CD20^+^CD27^+^IgD^+^), naïve B cells (CD19^+^CD20^+^CD27^-^IgD^+^), and double negative (DN) B cells (CD19^+^CD20^+^CD27^-^IgD^-^). The frequencies of these populations in the control group were similar to previously reported findings^19^, whereas the MS patients had reconstituted a predominantly naïve repertoire during RTX treatment interruption (Figure 4C).

**Figure 4:**
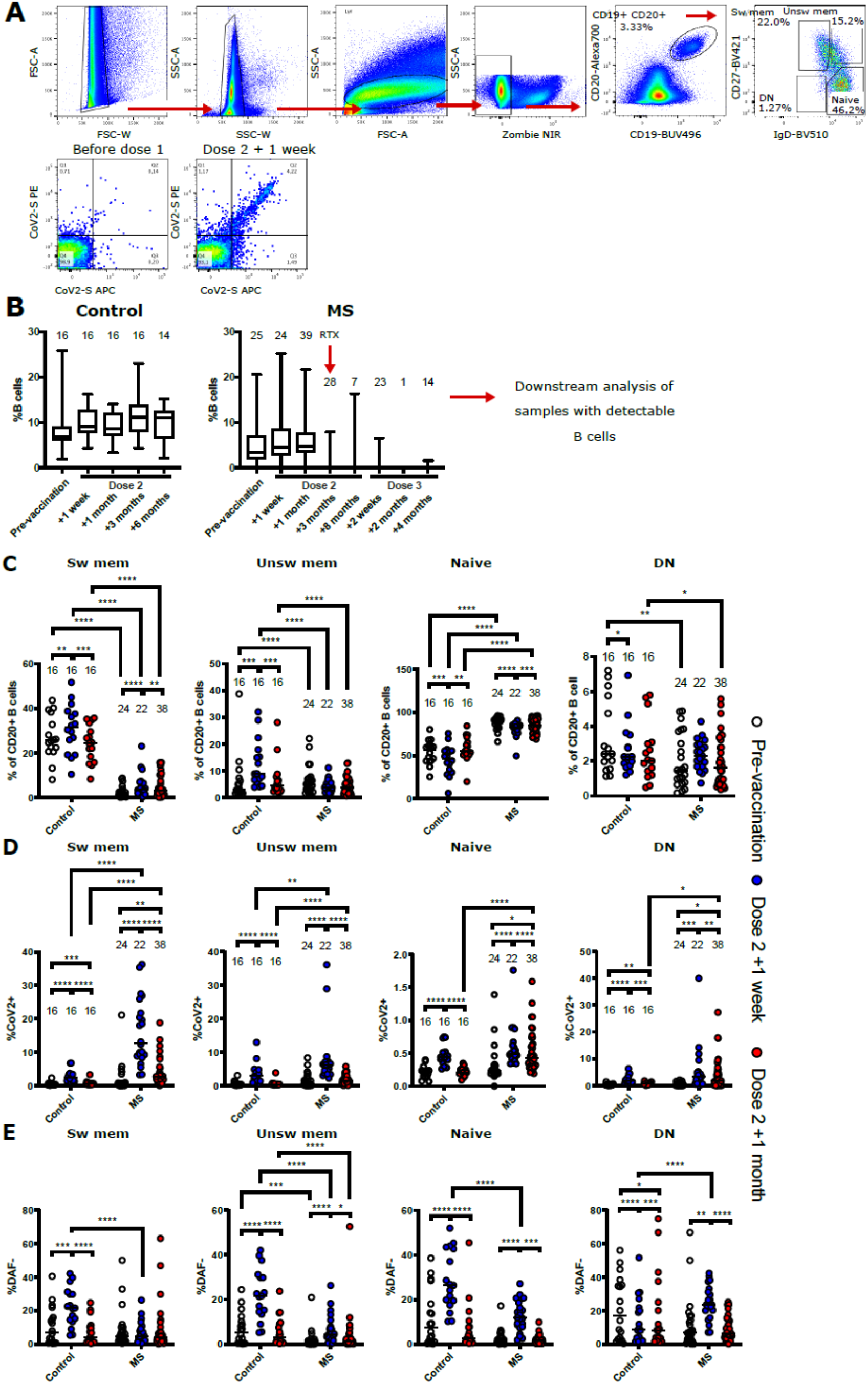
Flow cytometric characterisation of B cells from non-multiple sclerosis (MS) controls and MS patients. (A) Representative gating strategy for the final identification of switched memory (sw mem), unswitched memory (unsw mem), naïve, and double negative (DN) B cells. Definition of SARS-CoV2-specific B cells. (B) Longitudinal frequencies of B cells in non-MS controls and patients. Number of individuals are indicated above the respective box plots. Timepoint of re-initiated treatment with rituximab (RTX) is indicated in the patient group. (C-E) B cell frequencies in non-MS controls and patients before vaccination, one week and one month after the second vaccine dose. (C) Frequencies of sw mem, unsw mem, naïve, and DN B cells. (D) Frequencies of SARS-CoV2 specific B cells within the indicated subpopulations. (E) Frequencies of DAF-cells of each B cell subtype. * p<0.05; ** p<0.01; *** p<0.001; **** p<0.0001.

We further dissected the peripheral B cell compartment based on B cell expression of an S-specific B cell receptor (BCR). This was done to determine if prior RTX treatment had an impact on vaccine-induced responses in patients. We found significantly elevated levels of S-specific switched memory B cells in patients one week and one month after the second BNT162b2 dose, as compared to healthy controls at the same timepoints (Figure 4D). A similar difference was noted for S-specific unswitched memory B cells and we could also demonstrate that patients had a higher frequency of antigen-specific DN B cells than healthy controls at four weeks post a second BNT162b2 dose (Figure 4D). We have previously shown that B cells downregulate the complement regulatory protein DAF upon BCR engagement, and that DN B cells in circulation may reflect ongoing B cell responses in lymphoid organs^19,20^. We found that patients had increased frequencies of DAF^-^ cells in all B cell populations except for switched memory B cells one week after the second dose (Figure 4E). In comparison, the control individuals demonstrated increased levels of DAF^-^ B cells in all populations except DN B cells.

### SARS-CoV-2 specific DN B cell levels one month after the second vaccine dose correlate with high antibody response and virus neutralization after the third dose

We next did a quantitative correlation analysis of the serological and clinical parameters, as well as frequencies of different B cell populations (Figure 5 and supplemental Figure 4). This was done to better understand if there were parameters that could predict the antibody response magnitude or quality after vaccination of COVID-naïve MS patients on RTX treatment interruption.

**Figure 5:**
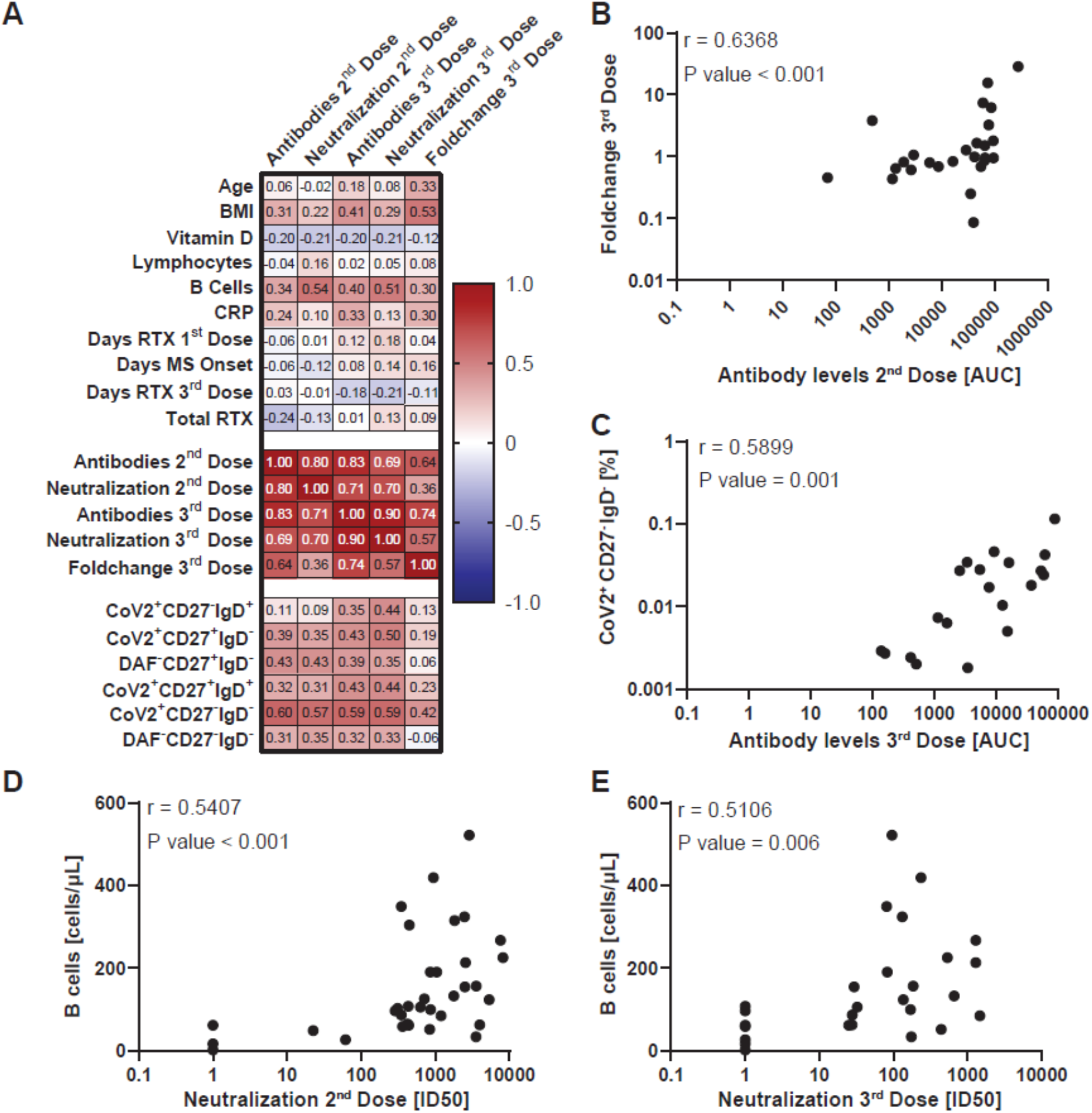
Correlation of vaccine response with demographic and clinical parameters in COVID-19 naïve multiple sclerosis (MS) patients. (A) Spearman correlation matrix between the SARS-CoV-2 spike-specific antibody levels together (AUC) with pseudotyped lentivirus neutralization capability (ID50) and multiple experimental and clinical parameters. Corresponding complete correlation matrix and P values are displayed in figures S3 and S4, respectively. Chosen parameters are listed in supplemental Method 3. (B-E) Spearman correlation of SARS-CoV-2 spike-specific antibody levels one week after the second vaccine dose and foldchange of SARS-CoV-2 spike-specific antibody levels after and before third vaccine dose (B), SARS-CoV-2 spike-specific antibody levels one week after the third vaccine dose and SARS-CoV-2^+^CD27^-^IgD^-^ B cell levels (C), total B cell levels before the first vaccine dose and SARS-CoV-2-pseudotyped lentivirus neutralization capability one week after the second (D) and third dose (E). The analysis results are displayed in the upper left.

We found that BMI, but no other demographic factors, had a strong correlation with the fold-increase of S-specific IgG before and after a third vaccine dose (Foldchange 3^rd^ Dose) (Figure 5A). Additionally, we found that levels of circulating anti-S IgG one week after a second dose correlated with the fold-increase after a third vaccine dose after re-initiation of RTX treatment (Figure 5B). Of the clinical parameters, the total B cell count before the first vaccination dose correlated with the neutralization capability after the second and third vaccine doses (Figure 5D,E). Out of all B cell populations, the SARS-CoV-2 spike-protein specific DN B cell frequencies one month after the second vaccine dose displayed the highest correlation with the antibody levels and neutralization capability after the second and third vaccine doses (Figure 5A,C). In summary, these findings add to the observation that circulating anti-S IgG after a second dose of BNT162b2 has predictive value for the boost effect after RTX treatment re-initiation of MS patients.

## Discussion

In this study we investigated humoral immune responses to SARS-CoV-2 vaccination in RTX-treated MS patients during interruption and subsequent re-initiation of treatment. We observed that MS patients with previous COVID infection had a strong response to both second and third vaccine doses, comparable to the non-MS control group. These data demonstrate that SARS-CoV-2 infection of MS patients on RTX treatment interruption generates a strong immunological memory that can be re-stimulated by vaccination, similarly to healthy individuals^21^. However, COVID-naïve MS patients displayed high variability in antibody levels after both second and third vaccine doses, showing similar or lower levels as COVID-experienced MS patients and non-MS controls.

Through B cell phenotype characterization, we could show that MS patients exhibited a significantly higher naïve B cell population and consequently lower memory (CD27^+^) subsets than the control group before and after SARS-CoV-2 vaccination. These findings coincide with previous reports of B cell reconstitution after RTX treatment, where the B cell populations were limited to immature, transitional, and mature naïve B cells, with low levels of memory subsets^22,23^. We additionally show that both MS patients and controls developed a robust SARS-CoV-2 S-specific memory B cell population upon vaccination. Although naïve B cell population frequencies were reported to be a general predictor for SARS-CoV-2 vaccine response in immunocompromised individuals^24^, this parameter alone could not explain the highly variable vaccine response of our COVID-naïve MS patients. Strikingly, SARS-CoV-2^+^ DN B cell frequencies were consistently higher in MS patients than in controls after vaccination. We show that this B cell subset correlates significantly with the antibody and neutralization levels after the second and third vaccine dose. To our knowledge, there has been no report investigating the function of DN B cells in RTX-treated MS patients. This B cell subset is inflated in patients with different chronic inflammatory conditions and after vaccination against influenza and tick-borne encephalitis virus^25,26^. In addition, DN B cells are shown to transcriptionally cluster with naïve, memory B cells, and plasmablasts, suggesting an extra-follicular maturation pathway^26^.

Germinal centre (GC) B cells express low levels of DAF, a phenomenon that we also observed in circulating B cells during acute Hantavirus infection^20^. We suppose that the increase of DAF^-^ B cells might reflect a robust GC response, since GC B cells downregulate their DAF expression. Indeed, the complement regulation in GCs has a crucial role in positive B cell selection and therefore antigen-induced antibody production^27^. Another factor that may be indicative of the GC activity is the avidity of elicited antibodies. Here, we found that MS patients had an overall lower avidity compared to the control group. The lower antibody avidity in MS patients in combination with the decreased expansion of DAF^-^ sw mem B cells might suggest that the B cell affinity maturation in the GC could be impaired as compared to the control group. Previous reports show that GC activity after RTX is negatively affected^28,29^. To date, however, there are no long-term effect studies on the impact of RTX on GC formation in MS individuals after treatment interruption. Our T cell analysis (supplemental Figure 4) showed a comparable trend in T cell populations between the control and patient groups after vaccination. Consistently, previous vaccination studies indicate the preservation of T cell levels and responses in anti-CD20 treated MS individuals^30^.

In summary, our findings highlight the heterogeneity of humoral immune responses to SARS-CoV-2 mRNA vaccination in RTX-treated MS patients and provide factors predictive of the response. The high vaccine response in a fraction of COVID-naïve MS patients hints towards the survival of SARS-CoV-2-specific memory B cells after re-initiation of RTX treatment. This may be due to differential depletion of different B cell populations within secondary lymphoid organs by RTX^29^. In addition, we show that SARS-CoV-2^+^ DN B cell frequencies one month after the second vaccine dose significantly correlate with a higher antibody response to the second and third vaccine doses. Importantly, S-binding antibody levels after the second vaccine dose could predict efficient immune responses to subsequent vaccine doses during ongoing RTX treatment. Taken together, this could aid the decision to antedate the third vaccine dose by several months while postponing re-initiation of RTX for individuals with poor immune responses to the first two vaccine doses. Lastly, our findings provide possible leads to research crucial aspects of vaccine responses in RTX-treated MS patients. The investigation of DN B cells and their potential extra-follicular maturation could provide deeper insight into alternative antigenic stimulation of B cells in these patients. Furthermore, the possible long-term impairment of GC activity in B-cell depleted MS patients needs further examination to understand and counteract their attenuated vaccine response.

## Data Availability

All data produced in the present work are contained in the manuscript

## Acknowledgements

The study nurses Anneli Sundström, Carina Olofsson, and Monica Holmgren are acknowledged for assisting in the study procedures. The laboratory technicians Maj Järner, Linnea Wikström for blood sample preparation. The study was supported by grants from Umeå University and Region Västerbotten (RV-969133), Swedish Research Council (Dnr 2020-0625 to M.NE.F. and Dnr 2021-04665), and the SciLifeLab National COVID-19 Research Program (VC-2020-0015) to M.NE.F..

## Authorship csontributions

R.G.: Designed and performed experiments, conducted analyses, and wrote the manuscript.

A.D.: Designed, performed, and analysed flow cytometric experiments, critically read and wrote the manuscript.

P.S., M.NE.F., C.A. and J.N.: Planned/conceived and designed the study, reviewed and edited the manuscript. PS: Responsible for recruitment of MS patients. C.A. and J.N. are principal investigators for the clinical trial and supervised sample and data collection.

## Methods

### Supplemental Method 1. Blood sampling numbers and intervals

In case of the rituximab-treated (RTX) multiple sclerosis (MS) group, blood samples were collected as depicted in supplemental Figure 1: from 24 individuals before the first vaccine dose, 23 at one week after the second, 40 at one month after the second, 28 at two to three months after the second, 20 at four to eight months after the second, 31 at one to two weeks after the third, 20 at one to two months after the third, and 14 at three to four months after the third. Two individuals, which had COVID-19 before vaccination, got only one vaccine dose. Their sampling timepoints after the first dose were defined as after second dose, in order to compare them with the remaining individuals. Regarding the non-MS control cohort, blood samples were collected from 19 individuals before the first vaccine dose, 19 at one week after the second, 19 at one month after the second, 17 at two to three months after the second, and 15 at four to eight months after the second.

For the avidity assay, blood samples for the control group were collected from 10 individuals at one week after the second vaccine dose, two to three months after the second, four to eight months after the second, and one to two months after the third. Samples for the RTX-MS group were collected from 10 individuals as a subgroup of the 43 individuals examined throughout the rest of the study. For this group, 10 samples were collected at one week after the second vaccine dose, two to three months after the second, one to two months after the third, and 5 samples at four to eight months after the second.

### Supplemental Method 2. SARS-CoV-2-pseudotyped lentivirus production

Approximately 19.5 million HEK 293T cells (CRL-3216 ATCC) were seeded per T175 flask in Dulbecco’s Modified Eagle Medium (DMEM) (Thermo Scientific) + 10 % fetal bovine serum (FBS) + 1000 U/L Penicillin+Streptomycin (D10 medium) for SARS-CoV-2-pseudotyped lentivirus production and incubated at 37 °C and 5 % CO2. On the next day, 109 µL of Fugene 6 (Promega) and 1100 µL of Opti-MEM (Thermo Scientific) per T175 flask was mixed in a sterile polystyrene tube and incubated for 5 min at room temperature (RT). 21.1 µg of pHR’ CMV Luc (luciferase reporter gene), 21.1 µg of pCMV delta8.2 (lentivirus backbone), 0.37 µg of TMPRSS2 (this and all previous plasmids were provided by Nicole Doria-Rose), and 1.2 µg of pHDM SARS-CoV-2 Spike D614G C-terminal deletion plasmid DNA (provided by Jesse Bloom) was added per T175 flask to the previous mixture and incubated for 15 min at RT. Finally, the reagent-DNA mixture was added to the media in the T175 flask containing the 293T cells and the flask returned to the incubator.

On the subsequent day, the medium in the T175 flask was exchanged with 19 mL of fresh D10 medium per flask and the flask returned to the incubator. On the following day, the cell supernatant was harvested in 50 mL tubes and centrifuged at 210xg for 5 min and passed through a 0.45 µm PVDF sterile filter (Millipore). Finally, the supernatant was aliquoted in 1 mL and stored at -80 °C.

### Supplemental Method 3. Parameters chosen for Spearman correlation

AUC of SARS-CoV-2 spike-specific antibody levels one week after the second vaccine dose, ID50 SARS-CoV-2-pseudotyped lentivirus neutralization capability one week after the second vaccine dose, SARS-CoV-2 spike-specific antibody levels one week after the third vaccine dose, SARS-CoV-2-pseudotyped lentivirus neutralization capability one week after the third vaccine dose, and foldchange of SARS-CoV-2 spike-specific antibody levels after and before third vaccine dose, days since last RTX treatment before first vaccine dose, days since MS disease onset before first vaccine dose, days since last RTX treatment before third vaccine dose, total administered RTX amount since treatment start [mg], blood vitamin D concentration before first vaccine dose [nmol/L], blood total lymphocyte count before first vaccine dose [10^9 cells/L], blood C-reactive protein levels before first vaccine dose [mg/mL], blood total B cell count before first vaccine dose [cells/µL], age, Body Mass Index, and frequencies of B cell populations [%] one month after the second vaccine dose (CD27^+^IgD^-^, SARS-CoV-2^+^CD27^+^IgD^-^, CD27^-^IgD^-^, SARS-CoV-2^+^CD27^-^IgD^-^).

## Tables

**Supplemental Table 1.** List of flow cytometric antibodies used.

| Antigen | Fluorophore | Clone | Vendor |
| --- | --- | --- | --- |
| CD19 | BUV496 | SJ25C1 | BD |
| CD20 | AlexaFluor-700 | 2H7 | Biolegend |
| CD27 | BV421 | MT271 | BD |
| IgD | BV510 | IA6-2 | BD |
| DAF | PE-Cy7 | JS11 | Biolegend |
| CD3 | BV510 | SK7 | Biolegend |
| CD4 | BUV395 | SK3 | BD |
| CCR7 | BV421 | 2-21-A | BD |
| CD45RA | BV786 | HI100 | BD |
| CD25 | BV711 | 2A3 | BD |
| DAF | APC | JS11 | Biolegend |

## Figures

**Supplemental Figure 1:**
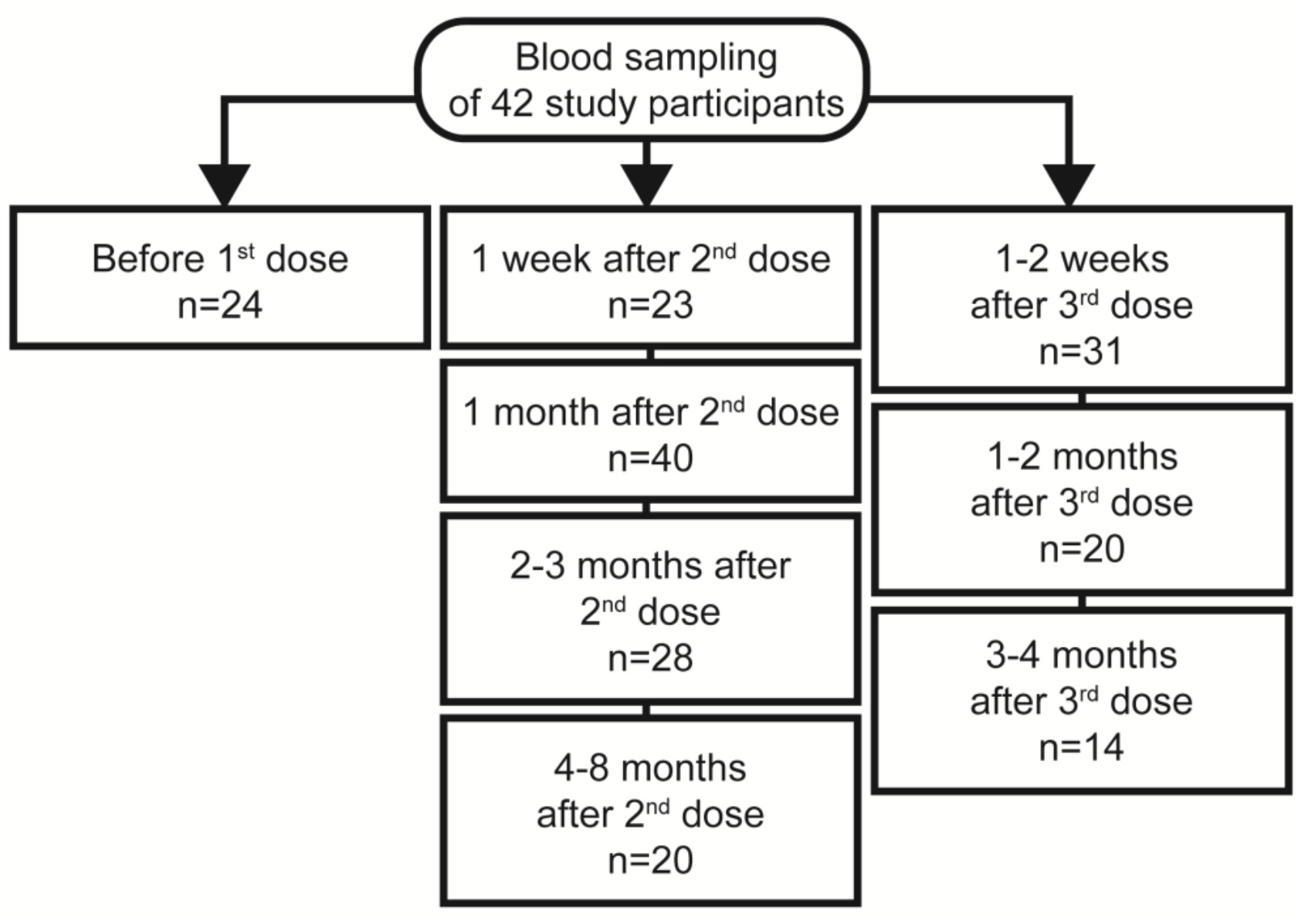
Blood sample time points and sample sizes of rituximab (RTX)-treated multiple sclerosis (MS) patients during SARS-CoV-2 vaccination.

**Supplemental Figure 2:**
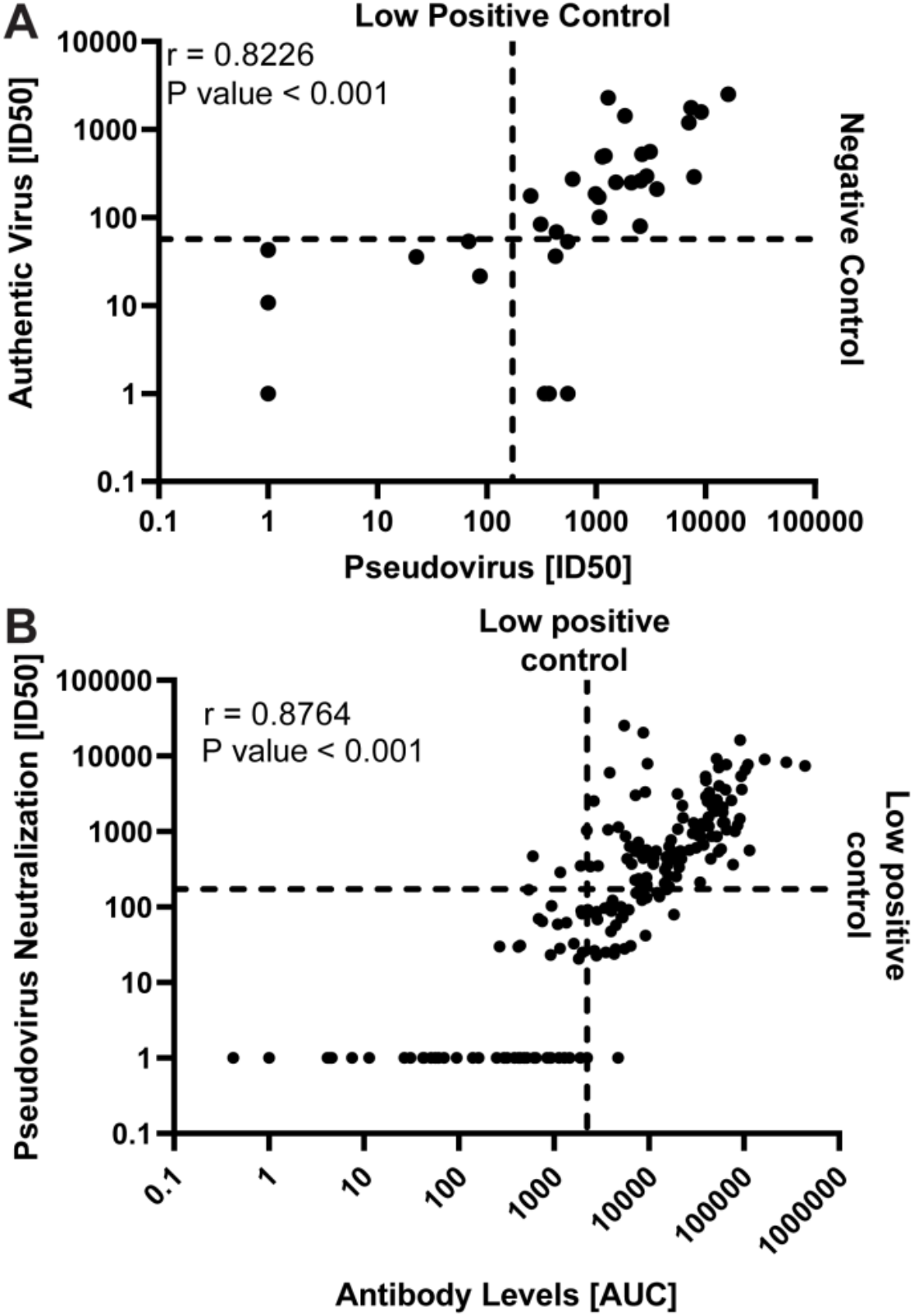
Correlation of authentic and pseudotyped SARS-CoV-2 virus neutralization. Comparison of authentic and pseudotyped SARS-CoV-2 virus neutralization assay (A), as well as SARS-CoV-2 spike-specific antibody levels and pseudotyped virus neutralization capability (B) of MS patient samples. Sample points, whose neutralizing capability or antibody levels were below detection have been set to 1 for illustrative purposes. Statistical parameters in the upper left describe the results of a Spearman correlation analysis. (A) Plot of authentic SARS-CoV-2 virus against SARS-CoV-2-pseudotyped lentivirus neutralization capability, described as the serum dilution required to reduce the pseudovirus infection by 50% (ID50), for MS patient serum samples one month after the second vaccine dose. The vertical and horizontal lines display the reference values for a low neutralizing and negative control serum sample, respectively. (B) Plot of SARS-CoV-2 spike-specific antibody levels (AUC) against SARS-CoV-2-pseudotyped lentivirus neutralization capability (ID50) for MS patient samples. The vertical and horizontal lines display the reference value for a low neutralizing control serum sample.

**Supplemental Figure 3:**
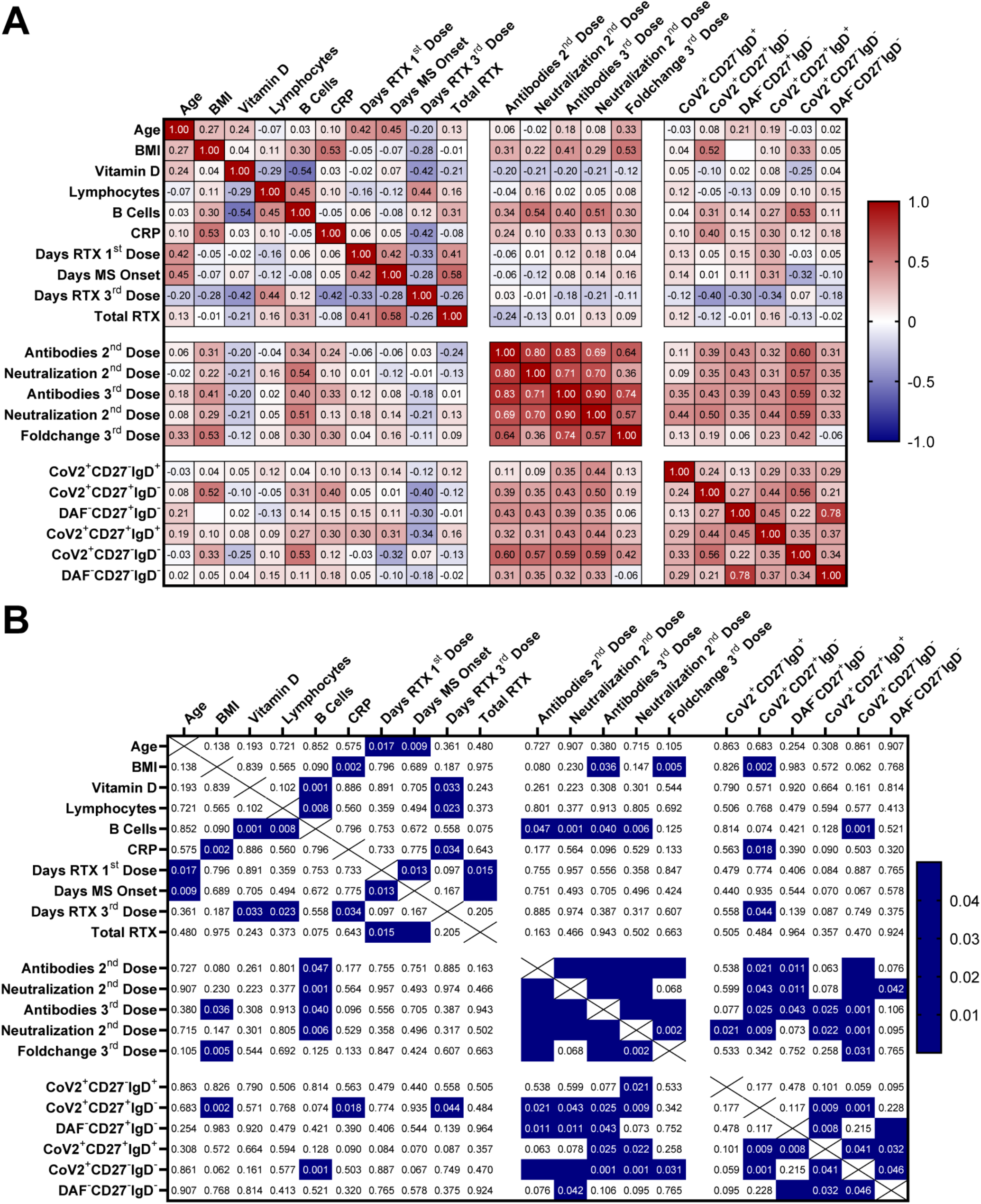
Frequencies of T cell populations based on their expression of CCR7 and CD45RA in non-multiple sclerosis (MS) controls and MS patients. Displayed are naïve (CCR7^+^CD45RA^+^), central memory (CCR7^+^ CD45RA^-^), effector memory (CCR7^-^ CD45RA^-^), and terminally differentiated effectors (CCR7^-^ CD45RA^+^). Upper row: CD4^+^ T cells. Middle row: CD8^+^ T cells. Bottom row: %CD25^+^ naïve and non-naïve CD4^+^ T cells.

**Supplemental Figure 4:**
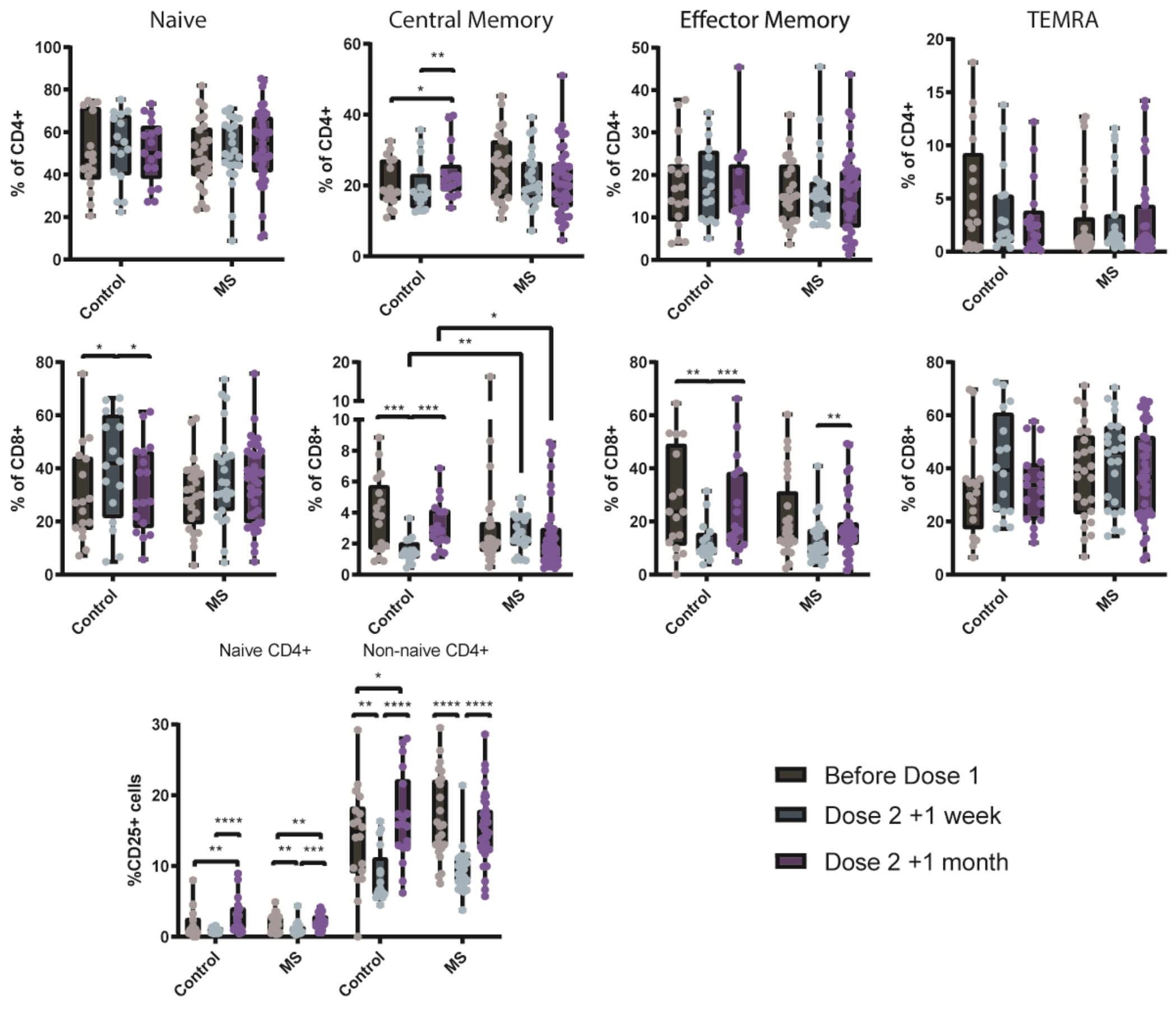
Complete correlation of multiple experimental and clinical parameters in COVID-19 naïve multiple sclerosis (MS) patients. (A) Spearman correlation matrix between multiple experimental and clinical parameters of COVID-19 naïve MS patients. (B) P values of the corresponding spearman correlation matrix. Matrix cells with a P value below 0.05 were depicted in blue. Chosen parameters are listed in the data analysis part of the materials and methods section.

